# Quantification of joint mobility limitation in adult type 1 diabetes

**DOI:** 10.1101/2023.05.08.23289681

**Authors:** Sanat Phatak, Pranav Mahadevkar, Kaustubh Chaudhari, Shreya Chakladar, Swasti Jain, Smita Dhadge, Sarita Jadhav, Rucha Wagh, Rohan Shah, Aboli Bhalerao, Anupama Patil, Jennifer Ingram, Pranay Goel, Chittaranjan Yajnik

## Abstract

**Background:** Diabetic cheiroarthropathies consist of limited joint mobility (LJM), flexor tenosynovitis (FTS), Dupuytren’s contracture (DC), and carpal tunnel syndrome (CTS). There is heterogeneity in definitions and lack of a method to measure hand fibrosis load. We measured metacarpophalangeal (MCP) joint restriction and describe magnetic resonance (MR) imaging characteristics across the spectrum of joint restriction.

**Methods:** Adults with type 1 diabetes were screened for hand manifestations using a symptom questionnaire, clinical examination, function (Duruoz hand index (DHI), grip strength). We measured maximum possible extension at the MCP joint. Patients were segregated by mean MCP extension (<20 degrees, 20-40 degrees, 40-60 degrees, and >60 degrees) for MRI scanning. Patients in the four groups were compared using ANOVA for clinical features as well as MRI measurements (tenosynovial, skin, and fascia thickness, additive score of three).

**Findings:** Of 237 patients (90 males), 79 (33.8%) had cheiroarthropathy; these had MCP extension limitation (39 degrees versus 61 degrees, p<0.01). Groups with restricted MCP extension were older, had higher prevalence of retinopathy and nephropathy, and higher DHI (1.9 vs 0.2) but very few (7%) had pain. MRI scans of the hand (n=61) showed flexor tenosynovitis in four and median neuritis in one. Groups with maximum MCP limitation had the thickest palmar skin but mean tendon thickness or median nerve area did not differ. The additive score could differentiate between levels of joint mobility restriction. Only mean palmar skin thickness was associated with MCP extension angle in multiple linear regression.

**Interpretation:** Joint mobility limitation, quantified by restricted MCP extension, was driven by skin thickening. MCP extension and fibrosis scoring on MRI can serve as quantitative measures of hand involvement for future associative studies.

## Introduction

The syndrome of limitation of joint mobility in type 1 diabetes was described by Rosenbloom in 1974.[1] Since then, the ‘diabetic cheiroarthropathies’, or musculoskeletal disorders of the hand in diabetes have expanded to include limited joint mobility (LJM), carpal tunnel syndrome (CTS), flexor tenosynovitis (FTS), and Dupuytren’s contracture (DC).[2, 3, 4] Often, these conditions may co-exist.[2] While these conditions may be seen in the healthy population, their prevalence is higher in diabetes.[5] A variable prevalence of 8-66% of LJM, 30% of CTS, 28% of FTS, and 9% of DC has been previously reported in patients with type 1 diabetes.[2, 6, 7, 8] This variability owes to a heterogeneity in definition, method of measurements, duration of diabetes in addition to population differences.

All the described entities occurring in the hand in diabetes are putatively fibrotic on histomorphology; biopsy studies have demonstrated excessive collagen deposition in periarticular connective tissue including tendon sheaths and increased collagen glycation.[9] Regardless of nomenclature, these conditions result in preferential inelasticity of structures on the palmar aspect of the hand, thus principally limiting finger and wrist extension. In severe cases, flexion contractures ensue, leading to the ‘prayer sign’, an inability to approximate the palms fully.[10] Severe cheiroarthropathy produces functional deficits, restricting patients’ ability to exercise effectively, thus indirectly adding to metabolic complications,[11] and affecting professional and self-care activities, causing financial burden and loss of independence.[12]

The presence of diabetic hand manifestations correlates with some, but not all, microvascular complications.[13] Irrespective of diabetes, hand tissue fibrosis, as seen in DC, is associated with fibrosis in internal organs, especially the liver and Peyronie’s disease.[14] Joint stiffness was also associated with higher blood pressure readings in the community.[15] Considering these reported associations, it is tempting to speculate that hand stiffness reflects a more global, profibrotic trajectory. Diabetes mellitus is associated with an increased risk of internal fibrosis, affecting the kidneys, heart, and liver, leading to morbidity and mortality.[16]

However, establishing such associations is currently hampered by the clinical heterogeneity, vagueness of definitions and lack of consensus in ‘measuring’ the severity of hand involvement: “limited joint mobility” and “prayer sign”, are at best, descriptive terms and do not convey the degree of fibrosis. To elucidate associations of hand manifestations with diabetes and organ fibrosis in more granular detail, quantitation of the amount of fibrosis in the hand, both in the clinic as well as using non-invasive imaging is required. An ideal clinical measurement should be able to encapsulate all the differing manifestations, and be easy and reproducible. While a few imaging descriptions of hand manifestations in diabetes exist,[17] no attempt has been reported to delineate structures involved nor measure the amount of fibrotic tissue in the hand in a systematic manner. To attempt such quantification, we conducted a clinical examination of the hands in a cohort of adult patients with type 1 diabetes. In a subset of patients chosen across a spectrum of levels of joint mobility limitation at the metacarpophalangeal (MCP) joints, we report magnetic resonance imaging (MRI) findings of the hand, including measurements of tissue thickness, and contributors to joint stiffness.

## Methods

### Patients and Clinical Setting

The study was conducted at the Diabetes unit, KEM Hospital Research Centre, a tertiary care specialized unit in Pune, India. We screened consecutive adult patients (>18 years of age) with type 1 diabetes attending the Type 1 diabetes clinic, from March 2021 to December 2022. Type 1 diabetes was initially classified clinically, viz diagnosed <30 years, ketosis at presentation, and clinical dependence on insulin treatment; later confirmed with C-peptide measurements and the presence of islet autoantibodies. We excluded pregnant patients, those who needed hospital admission, and those with previous hand trauma or concurrent inflammatory arthritis involving the hand joints. Demographic information including age, gender, residence, education, and occupation were recorded. Those who worked in agriculture, manual labor or operating heavy machinery for more than 4 hours a day were classified as manual work. Those who worked on computer keyboards for more than six hours a day were classified as keyboard workers. Smoking (current, previous, never) and alcohol habits were recorded.

Data concerning the course of diabetes included date of diagnosis, insulin compliance, and the use of alternative medications. Evidence of micro- and macro-vascular complications was extracted from patient files: including evidence of retinopathy in fundus photographs, nephropathy from the presence of proteinuria and/or end stage renal disease, neuropathy based on biothesiometry and clinical composite score Michigan Neuropathy Screening Instrument, and coronary artery disease. Common comorbidities, such as hypertension and hypothyroidism, were recorded; we also asked for conditions that could possibly contribute to profibrotic manifestations including epilepsy, systemic sclerosis, skin disease involving the palms or hands, and malignancy. Similarly, we asked for a history of treatments that could possibly be fibrogenic, including methotrexate, amiodarone, aspirin, statins, and anti-epileptics.

Anthropometric measurements (height and weight) and blood pressure were recorded using standard procedures. We also noted the presence of keloid scars and lipo-hypertrophy or atrophy at insulin injection sites. We used skin autofluorescence as an indirect measurement of advanced glycation end-product (AGE) measurements using the AGE reader, that has been previously linked to complication burden in type 1 diabetes.[18] The reader provides a quantitative estimate, as well as a risk category calculated considering age and duration of diabetes. These measurements were performed on the forearm of the non-dominant hand, in standardized light conditions.

### Quantitative measures of hand manifestations

We screened for hand involvement using a structured history, clinical examination, and measurements by trained research staff. The musculoskeletal history included presence of pain on the palmar surface of the hand, symptoms of compressive neuropathy (sensory and motor), grip difficulty, finger triggering, and perceived stiffness and tightness of palmar skin. Hand function was measured using the Duruoz Hand Index (DHI), a self-reported questionnaire that assesses activity limitation in 18 daily activities each measured on a visual analogue scale from 0-5 with possible scores ranging from zero to 90, that has been validated for use in diabetes related hand conditions.[19]

Hand examination included the prayer sign, defined as a visible gap between the two palms with an inability to approximate them fully. We measured the distance between the two 5th metacarpophalangeal (MCP) joints and 5th proximal interphalangeal (PIP) joints, viewed from the ulnar aspect.[Figure 1B] As a measure of limited joint mobility, we recorded maximum possible passive extension at the MCP joint (2nd to 5th of both hands) until restriction or pain, with the palm approximated on a flat surface, using a protractor.[Figure 1A] Mean passive MCP extension was calculated for each hand. The presence of flexor tendon thickening, nodularity, triggering, and crepitus was noted. For the presence of carpal tunnel syndrome, we examined for the Tinel sign in both hands, the Phalen sign and sensation in the median nerve distribution, recorded as normal, reduced, or absent. Hand grip strength was measured for both hands, using a Jamar hand dynamometer (Patterson Medical, Warrenville, IL) Three readings were taken for each hand and a mean was calculated.

**Figure 1:**
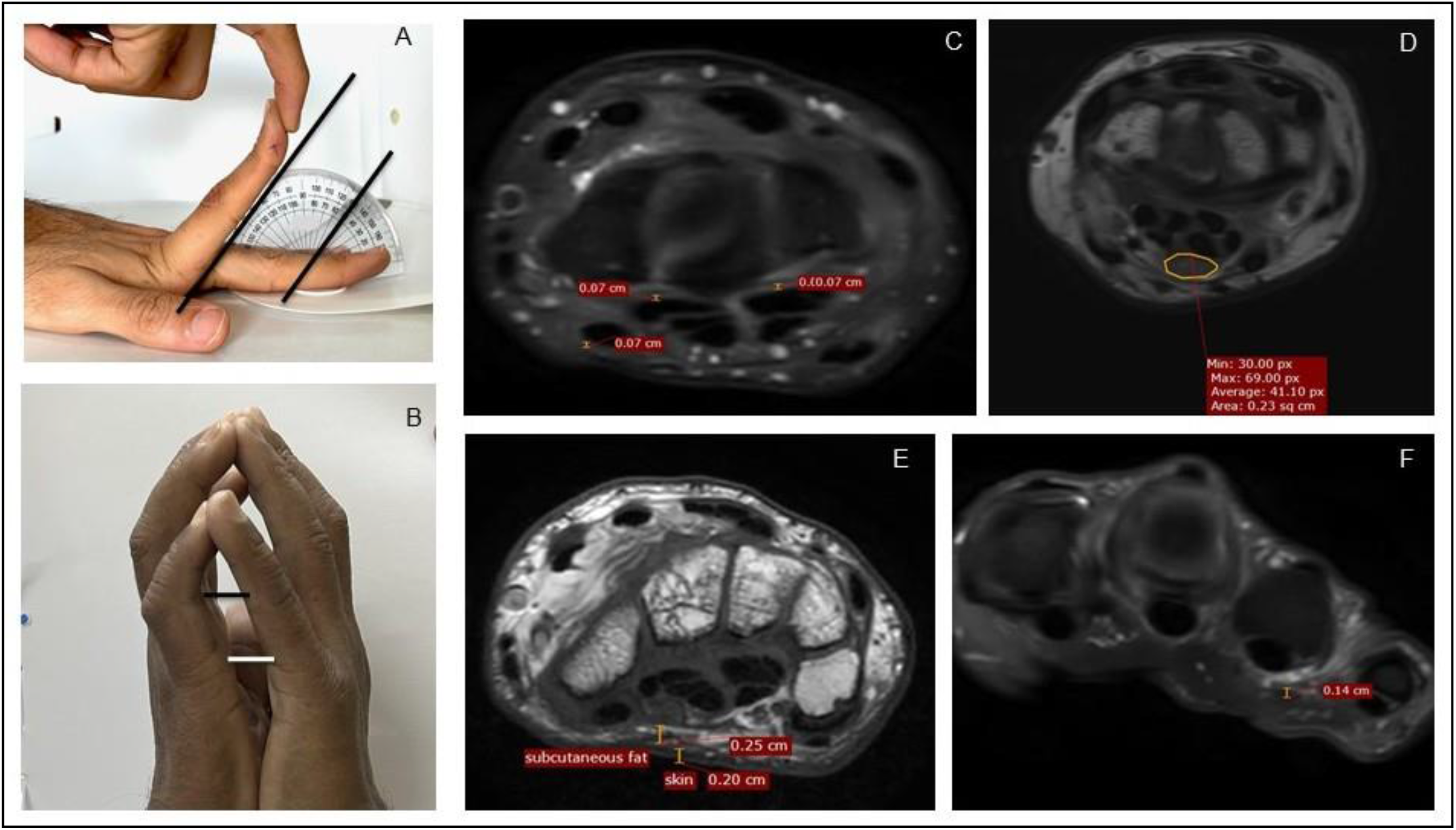
Measurement methodology, Clinical and MRI. A: measurement of maximum passive extension at MCP joint of left second finger, here showing 50 degrees extension. B: Measurements of distance between fifth proximal interphalangeal joints and fifth MCP joints as a quantification of the prayer sign. C: Measurement of tenosynovial thickness at the level of carpal tunnel; D. Measurement of median nerve area at the level of the carpal tunnel E. Measurement of skin and subcutaneous fat and F: Measurement of palmar fascia overlying the fourth flexor tendon on PDFS images. MRI: magnetic resonance imaging; PDFS: Proton density fat saturation

A physician (rheumatologist or diabetologist) examined all patients independently without access to the above measurements, and opined if one or more hand manifestation was present (LJM, flexor tenosynovitis/trigger finger, CTS, DD) was present, along with a physician-perceived severity (mild, moderate, severe). Inter-rater reliability between two physicians (SD, SP), seen in 30 patients, was 0.88.

### Selection of patients for MRI scans

Regardless of the cause of limited joint restriction, viz., LHM, DD or FT, or the number of fingers affected, we expected mean passive MCP extension to be compromised. Thus, non-dominant hand mean passive MCP extension was used as a metric of joint mobility restriction to segregate patients into 20-degree bins (0-20 degrees, most severe joint mobility limitation; 20-40 degrees, 40-60 degrees and more than 60 degrees).

Patients were approached for an MRI scan of the hand, all performed at the Star Imaging Research Centre using a 3T MRI Superconducting system with eight channel extremity coil (Ingenia Release 5, Philips Healthcare, Amsterdam, The Netherlands). Patients who agreed were invited to the Diabetes Unit, where earlier clinical findings were confirmed. Contraindications to MRI were excluded (metallic bone, cardiac, cochlear, or dental implants, metallic intrauterine contraceptive devices, pregnancy, and claustrophobia). Two patients were on insulin pumps, both the pump and sensor-transmitters were removed for the scan. Random plasma glucose measurements were performed, and the diabetologist (SD) managed insulin dosages accordingly, to prevent hypoglycemia whilst inside the MRI scanner. MRI of the non-dominant hand was performed, unless hand involvement was unilateral, in which case, the involved hand was imaged.

### MR Sequences and measurements

The following MR sequences were performed: Axial and coronal T2 weighted images, pre-contrast fat saturated T1-weighted axial coronal with fat-saturation. All MRI scans were read by the same musculoskeletal radiologist (PM) who provided a qualitative report (altered signals, thickening, edema) on the status of bones, tendons, joints, median nerve, and other salient findings. In addition, quantitative measurements were performed by one of two trained researchers (SC, SJ) on axial images, in addition to the radiologist (PM). The readers had good internal consistency (Cronbach alpha 0.95) and agreement with each other (correlation coefficient 0.78). Measurements included tenosynovial thickness for the four flexor digitorum longus tendons and the flexor hallucis longus tendon, at four levels: at the metacarpophalangeal joint, the midpoint of the proximal phalanx, midpoint of the metacarpal bone, and the midpoint of the carpal tunnel.[Figure 1C] Mean tenosynovial thickness was calculated as average of these 20 data points. In addition, the thickness at a point of visually perceived maximum thickness for each tendon sheath was recorded. Skin and subcutaneous tissue thickness was measured at 4 points at the level of MCP joint, and four points at the level of the carpal tunnel, and an average was calculated.[Figure 1E] Palmar fascia was visible only when thickened; it was measured in those who had nodular thickenings (DC) at the site of thickening and was considered zero in others.[Figure 1F] The median nerve cross sectional area was measured at the level of the carpal tunnel outlet and median nerve signal abnormalities were noted.[Figure 1D] An instinctive ‘total hand fibrosis’ score was calculated by adding mean tenosynovial thickness, palmar fascia thickness and palmar skin thickness. A similar score was also calculated using Z-scores of mean tenosynovial thickness, palmar fascia thickness, and palmar skin thickness (total hand fibrosis Z-score). Patients with least joint flexibility had highest skin thickness on hand MRI.[Figure 2]

**Figure 2:**
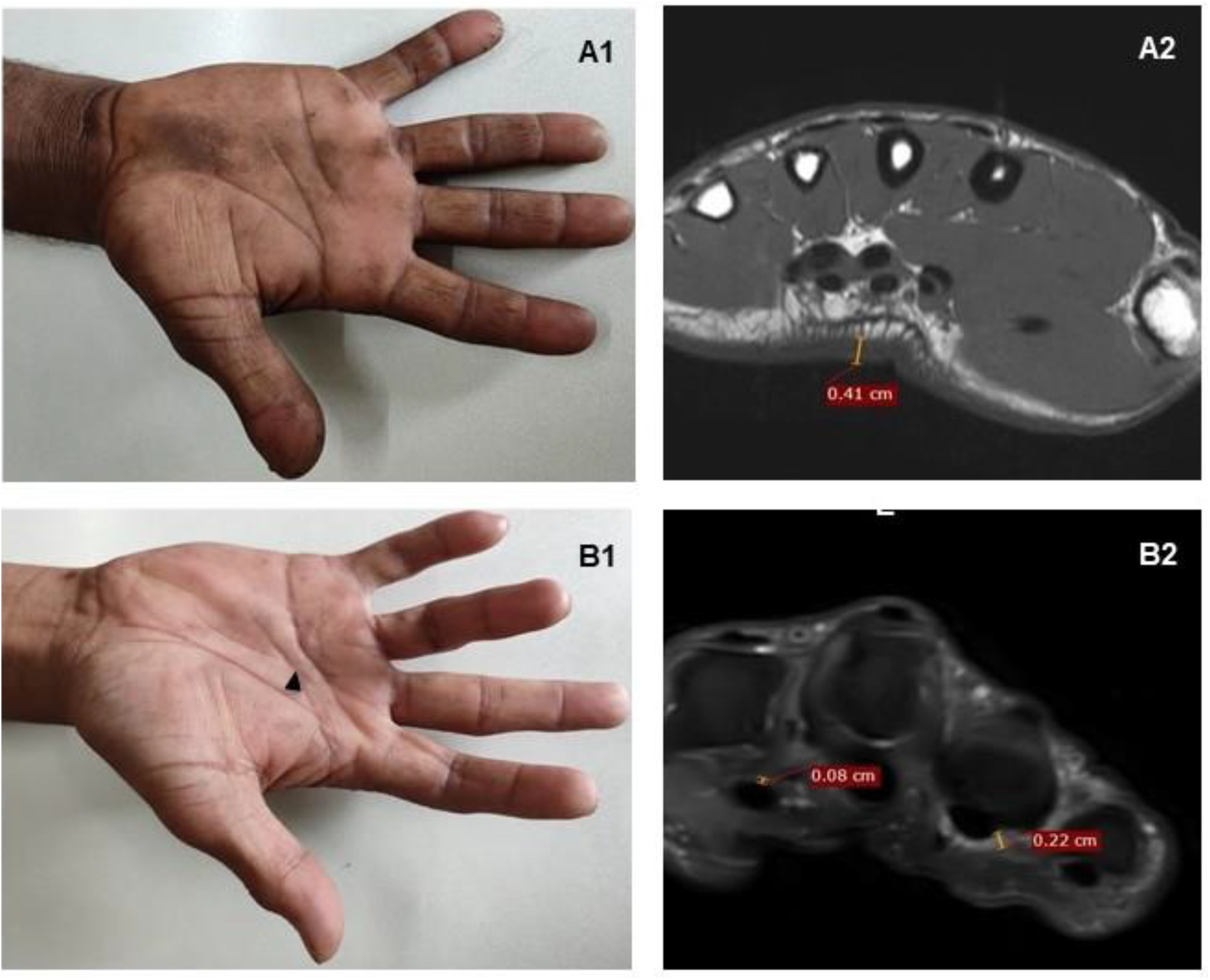
MRI findings in two patients with MCP extension restriction. A1: A patient with severely restricted MCP extension – mean 10 degrees. A2: MRI demonstrates considerable palmar skin thickening alone on T1 weighted axial images, seen at mid-metacarpal level; there is no tenosynovial or palmar fascia thickening. B1: A patient with bilateral Dupuytren’s contracture on the fourth finger (black arrow); PDFS scan shows Ill-defined PDFS hyperintense soft tissue thickening in relation to flexor thickening also involving A1 pulley and the palmar fascia.

### Statistical analysis

Data are presented as mean (standard deviation) and median (interquartile range) as appropriate for the variable. Patients who were judged to have diabetic cheiroarthropathy were compared with those without, using t-test. Patients with different levels of MCP extension limitation in 20-degree bins (0-20 degrees, 20-40 degrees, 40-60 degrees, and >60 degrees) were compared with each other with respect to demographics, diabetes characteristics and MRI characteristics using ANOVA; we report p value for trend, with <0.05 considered significant. MRI quantitative descriptors are described as mean (standard deviation) for each group. Multiple linear regression was used for structural determinants of hand stiffness: we used average MCP angle of the imaged hand as the dependent variable, while average skin thickness, average subcutaneous thickness, tenosynovial thickness, palmar fascia thickness, and median nerve cross-sectional area were used as predictor variables. We first standardized the independent and the dependent variables of the dataset into their corresponding Z-scores. All statistical analysis were performed using SPSS (IBM corporation, Armonk, NY) and R (R Foundation, Vienna, Austria).

#### Ethics

This study received ethics permission from the KEM Hospital Research Centre Ethics Committee (KEMHRC/RVC/EC/1518) and patients signed separate informed consent forms for clinical examination, photographs, and MRI scan. The study was registered with the Clinical Trials Registry of India (CTRI/2020/12/030057). Data sharing agreements were signed with Star Imaging and Research Centre and Indian Institute of Science Education and Research (IISER), Pune. The study received a waiver from the IISER Ethics Committee for Human Research (IECHR, Admin/2021/007). All clinical and imaging data are stored at the Diabetes Unit, KEM Hospital Research Centre.

#### Role of Funding

This study is funded through a DBT/Wellcome India Alliance Clinical and Public health fellowship (IA/CPHE/19/504607). The funding body had no role in study design or analysis.

## Results

### Cohort characteristics [Table 1]

We examined 237 adults with type 1 diabetes (90 males, median age 26.8 years). Most were college students, 15 (6%) operated heavy machinery, and 3 worked on keyboards for more than 6 hours. All had received their diagnosis of diabetes in childhood and had median duration of diabetes of 13.7 years. All took injectable insulin, and 57 also received metformin. Two patients used insulin pump. One-fifth of the cohort (20 patients, 8.5%) had retinopathy, 28 (11.8%) had nephropathy, and 45 (19%) had neuropathy on clinical examination.

**Table 1:** Patient Characteristics

| Patient Characteristics | n = 237 |
| --- | --- |
| Male | 90 (40.9%) |
| Age in years (median, range) | 26.8 (20.4, 36.0) |
| Age At Diagnosis, years (median, range) | 12.3 (9.2, 16.4) |
| Duration of diabetes (median, range) | 13.7 (8.1, 22.4) |
| Occupation: manual | 15.0 (6.3%) |
| Occupation: Keyboard >6 hrs | 3.0 (1.3%) |
| Nicotine use, Current or past | 21.0 (8.9%) |
| Alcohol use, current or past | 25.0 (10.5%) |
| Metformin use (n%) | 57.0 (24.0%) |
| Retinopathy | 20 (8.4%) |
| Proliferative | 7 |
| Nonproliferative retinopathy | 13 |
| Nephropathy (n%) | 28 (11.8%) |
| Neuropathy, clinical | 45 (18.9%) |
| On NCV (n%) | 1 (2.2%) |
| Presence of additional fibrogenic disease |  |
| Exposure to possible fibrogenic medication | 34.0 (14.3%) |
| Weight (kg) (median, range) | 56.8 (49.0, 66.1) |
| Height (cm, median, range) | 160.0 (154.0, 169.0) |
| BMI (kg/cm<sup>2</sup>) (median, range) | 22.0 (19.1, 24.9) |
| Lipohypertrophy | 49 (20.7%) |
| Lipoatrophy | 3 (1.3%) |
| Keloid scar | 14 (5.9%) |
| AGE reading (median, IQR) | 2.4 (1.9, 2.9) |
| Hand pain on the day of assessment | 9 (3.8%) |
| Finger locking/ triggering, current or past | 26 (10.9%) |
| Paresthesia in hands, current or past | 16 (6.8%) |
| Frozen shoulder |  |
| Duruoz Hand Index, median | 0 |
| Duruoz hand index >10 n, % | 6 (2.5%) |
| Grip strength, dominant hand (mm Hg), median, IQR | 41.7 (33.3, 57.5) |
| Grip strength, non-dominant hand (mm Hg), median, IQR | 41.7 (33.3, 56.7) |
| Prayer sign (n%) | 73.0 (30.8%) |
| Distance between 5th MCP, cm (mean, SD) | 0.815 (3.01) |
| Distance between 5<sup>th</sup> PIP, cm (Mean SD) | 1.116 (3.10) |
| MCP extension angle, degrees - Dominant hand (mean SD) | 50.8 (16.7) |
| MCP extension angle, degrees - Non-dominant hand (mean SD) | 54.2 (16.9) |
| CTS (Tinel sign or Phalen sign or reduced/absent sensation) | 53 (22.4%) |
| Physician diagnosis of diabetic cheiroarthropathy (n%) | 79 (33.8%) |
| Dupuytren's disease (n%) | 3 (1.2%) |
| Flexor tenosynovitis (n%) | 30.0 (12.6%) |
| Limited joint mobility (n%) | 46.0 (19.4%) |
| CTS (n%) | 24 (10.1%) |
| >1 condition | 24 (10.1%) |

### Musculoskeletal history and hand examination

Only nine (3.8%) complained of hand pain at the time of clinical assessment; 26 had a history of trigger finger. Sixteen (6.8%) complained of paresthesia and numbness in the hands. As a group, mean passive extension at the MCP joint was 50.8 degrees (SD) on the right hand and 54.2 degrees on the left. Prayer sign was seen in 73 (30%). Fifty-three had carpal tunnel syndrome on clinical examination. Seventy-nine (33.8%) had a cheiroarthropathy manifestation on physical examination; 30 had flexor tenosynovitis, 46 had LJM, 24 had CTS, and three had DC. Twenty-four had more than one condition. Patients with a physician-diagnosed cheiroarthropathy had a significantly different average MCP extension angle (39 degrees vs 61 degrees, p<0.01) and a significantly higher DHI (1.67 vs 0.21, p<0.01).

### Patient characteristics as divided by MCP extension limitation

Average MCP extension (2nd to 5th finger) of the non-dominant hand had a range of zero to 88 degrees; patients were grouped into four bins of 20 degrees each.[Table 2] Most patients (103, 43%) had mean extension in the 40-60 degrees range, while 64 (27%) had extension limited to less than 40 degrees. The group with the most severe limitation (<20 degrees) was predominantly male (11/15, 73%), while all the other groups had higher numbers of female participants. Groups with restriction (0-20, 20-40) tended to be older than the groups without and had diabetes for nearly a decade longer. One-fifth of the <20 degrees group had participated in regular manual labor; the group also had a much higher prevalence of smoking and/or alcohol use (nearly 30%) as compared to the other groups (nearly 10%). The group with the most severe restriction also had the highest prevalence of retinopathy (20%) and nephropathy (33%), but not neuropathy. Body composition did not vary across groups of hand stiffness; subjects with MCP restriction had a small but statistically significant difference in systolic blood pressure but not diastolic. They also had the highest prevalence of lipohypertrophy and keloid scar formation. Autofluorescence on AGE reader did not differ in the four groups.

**Table 2:** Clinical findings in patients grouped by degree of joint mobility restriction

| **Average MCP extension angle, nondominant hand categories (degrees)** | **0-20** | **20-40** | **40-60** | **>60** | **P for trend** |
| --- | --- | --- | --- | --- | --- |
| No | 15 | 49 | 103 | 69 |  |
| Male | 11 (73.3%) | 19.0 (38.8%) | 45 (43.7%) | 22 (31.9%) | 0.02 |
| Age, years (median, range) | 33 (26, 44) | 34 (23, 43.5) | 25 (19, 33) | 22 (19, 30) | 0.00 |
| Age At Diagnosis, years (median, range) | 13.5 (6.6, 14.4) | 12 (8.8, 16.9) | 12.4 (9.2, 15.9) | 11.8 (9.3, 16) | 0.51 |
| Duration of diabetes, years (median, range) | 23.9 (18.9, 33.6) | 22.1 (13.2, 30.3) | 11.4 (7.2, 18.7) | 10.4 (7, 15.8) | 0.00 |
| Occupation: manual labour | 3 (20.0%) | 6 (12.2%) | 4 (3.9%) | 2 (2.9%) |  |
| Keyboard >6 hrs | 0 | 0 | 2 (1.9%) | 1 (1.4%) |  |
| Current or past smoking (n, %) | 4 (26.7%) | 4 (8.2%) | 8 (7.8%) | 5 (7.2%) | 0.10 |
| Current or past alcohol (n, %) | 5 (33.3%) | 3 (6.1%) | 10 (9.7%) | 7 (10.1%) | 0.22 |
| Metformin use (n, %) | 5 (33.3%) | 12 (24.5%) | 20 (19.8%) | 19 (27.5%) | 0.50 |
| Retinopathy (n, %) | 3 (20.0%) | 7 (4.3%) | 7 (6.9%) | 5.0 (7.4%) | 0.06 |
| Nephropathy (n, %) | 5 (33.3%) | 3.0 (6.1%) | 14.0 (13.6%) | 6.0 (8.7%) | 0.11 |
| Neuropathy, clinical (n, %) | 2 (13.3%) | 15.0 (30.6%) | 19.0 (18.4%) | 9.0 (13.0%) | 0.11 |
| Exposure to fibrogenic medication | 5 (33.3%) | 15 (30.6%) | 10 (9.7%) | 3 (4.3%) | 0.00 |
| Weight (kg) (median, range) | 60.8 (44.6, 67.5) | 58.2 (52, 69.2) | 55.3 (49, 64.4) | 56.6 (48, 65.4) | 0.14 |
| Height (cm) (median, range) | 163 (150, 167) | 159 (154.3, 168.8) | 160.5 (153.4, 170) | 160 (153.8, 169) | 0.92 |
| BMI (kg/cm<sup>2</sup>) (median, range) | 21.9 (20.1, 24.7) | 23.1 (20.5, 26.3) | 21.5 (19, 24.8) | 21.7 (18.7, 24.9) | 0.05 |
| Systolic BP (mm Hg) (median, range) | 118 (110, 148) | 122.5 (111, 139.8) | 116 (109, 124.4) | 114 (104.3, 120) | 0.00 |
| Diastolic BP (mm Hg) (median, range) | 74.5 (67, 80) | 76.5 (68.3, 84.5) | 73.5 (67.3, 80) | 72.5 (68, 79.9) | 0.17 |
| Lipohypertrophy (n%) | 6 (40.0%) | 11.0 (22.4%) | 20.0 (19.4%) | 12.0 (17.4%) | 0.09 |
| Lipoatrophy (n%) | 0 | 0 | 2 (1.9%) | 1 (1.5%) | 0.53 |
| Keloid scar (n%) | 2 (13.3%) | 2.0 (4.1%) | 6 (5.8%) | 4 (5.8%) | 0.75 |
| AGE reading (median, IQR) | 2.4 (2.2, 2.7) | 2.6 (2.2, 3.1) | 2.2 (1.7, 2.8) | 2.5 (2, 2.9) | 0.17 |
| Hand pain on day of exam | 1 (6.7%) | 3 (6.1%) | 4 (3.9%) | 1 (1.5%) | 0.17 |
| Finger locking | 3 (20.%) | 12 (24.5%) | 7 (6.8%) | 4 (5.8%) | 0.00 |
| Paresthesia | 2.0 (13.3%) | 7.0 (14.3%) | 5.0 (4.8%) | 2.0 (3.0%) | 0.01 |
| Frozen shoulder | 3 (20.%) | 12 (24.5%) | 5 (4.8%) | 0 | 0.00 |
| Duruoz Hand Index Mean | 1.87 | 1.94 | 0.26 | 0.23 | 0.00 |
| Duruoz Hand Index >10 (n%) | 1.0 (6.7%) | 3.0 (6.1%) | 1.0 (0.97%) | 1.0 (1.5%) | 0.07 |
| Grip strength, dominant hand (mm Hg), median, IQR | 46.7 (36.7, 63.3) | 43.3 (32.5, 62.5) | 43.3 (33.3, 60.0) | 40.0 (32.5, 48.3) | 0.03 |
| Grip strength, non-dominant hand (mm Hg), median, IQR | 51.7 (40.0, 63.3) | 41.7 (33.3, 63.3) | 45.0 (33.3, 56.7) | 40.0 (31.7, 46.7) | 0.01 |
| Prayer sign (n%) | 12 (80.0%) | 24 (48.9%) | 10 (9.7%) | 27 (39.1%) | 0.00 |
| Distance between 5th MCP (cm) (mean, SD) | 3.14 (4.54) | 1.73 (5.77) | 0.43 (0.93) | 0.26 (0.66) | 0.00 |
| Distance between 5<sup>th</sup> PIP (cm) (Mean SD) | 5 (6.86) | 2.15 (4.69) | 0.64 (1.43) | 0.29 (0.79) | 0.00 |
| MCP extension angle, degrees - Dominant hand (mean, SD) | 13 (7.1) | 32.9 (5.2) | 53.4 (5.8) | 67.8 (5.5) | 0.00 |
| MCP extension angle, degrees - Non-dominant hand (mean, SD) | 16.2 (11.4) | 36.6 (7.1) | 57.4 (7) | 70.1 (6.4) | 0.00 |
| Physician diagnosis of diabetic cheiroarthropathy (n%) | 15 (100%) | 35 (71.4%) | 21.0 (20.4%) | 7.0 (10.1%) | 0.00 |
| **Number of MRI scans** | **12** | **14** | **20** | **15** | **P for trend** |
| Mean tendon sheath thickness (mm) | 0.58 (0.55, 0.61) | 0.55 (0.46, 0.64) | 0.5 (0.45, 0.55) | 0.65 (0.52, 0.7) | 0.821 |
| Maximum tendon sheath thickness (mm) | 0.7 (0.63, 0.88) | 0.64 (0.55, 0.77) | 0.56 (0.52, 0.63) | 0.72 (0.56, 0.82) | 0.1 |
| Mean skin thickness (mm) | 1.48 (1.1, 1.7) | 1.4 (1.03, 1.6) | 1.38 (1.09, 1.55) | 0.95 (0.8, 1.25) | 0.003 |
| Mean subcutaneous thickness (mm) | 2.58 (2.08, 3.48) | 2.48 (2.25, 3.16) | 2.15 (1.71, 2.74) | 3.5 (2.5, 4.3) | 0.211 |
| Mean median nerve area (mm<sup>2</sup>) | 8.95 (8.03, 9.9) | 10 (8.88, 11.3) | 8.6 (7.1, 9.1) | 9 (6.88, 10.2) | 0.076 |
| Total fibrosis: Sum mean tendon sheath, Skin thickness and palmar fascia thickness | 2.045 (1.718, 2.271) | 1.933 (1.577, 2.196) | 1.938 (1.649, 2.055) | 1.585 (1.445, 1.774) | 0.003 |
| Z-score: Sum Z-score of mean tendon sheath, palmar fascia, and skin thickness | 0.607 (-0.373, 1.187) | -0.236 (-1.164, 1.38) | -0.3 (-1.412, 0.331) | -0.354 (-1.126, 0.093) | 0.023 |
| Additive MRI fibrosis (Mean skin thickness + mean tendon thickness + mean palmar fascia thickness) | 2.40 (1.4) | 2.02 (0.7) | 1.87 (0.34) | 1.64 (0.30) | 0.011 |
| Z-score addition score | 1.32 (3.37) | 0.03 (1.65) | -0.48 (1.16) | -0.45 (1.1) | 0.018 |

Hand joint restriction was rarely painful: Only 7% complained of hand pain even in the most restricted group; one fourth had a history of trigger finger. Similarly, though more often seen in the restricted groups, only a small fraction had symptoms (14%) of neuropathy. Both observations translated into a small but statistically significant higher DHI in the restricted groups (mean 1.9 compared to 0.2). Only six percent in the <20 degrees group had a DHI of more than 10, that we considered clinically relevant hand function restriction. Most patients (80%) were observed to have a prayer sign in the <20 degrees group, and they consequently also had the maximum distance between the two MCP joints (mean 3 cm) and PIP joints (mean 5 cm). The examining physician in an independent opinion felt all patients in the <20 degrees group and three-fourth of the 20-40 degrees group had at least one described cheiroarthropathy phenotype. In univariate linear regression analyses adjusted for gender and duration of diabetes, height, systolic blood pressure, urine albumin creatinine ratio/clinically determined nephropathy, and the presence of frozen shoulder was significantly associated with average non-dominant hand MCP extension angle.[Table 3]

**Table 3:**
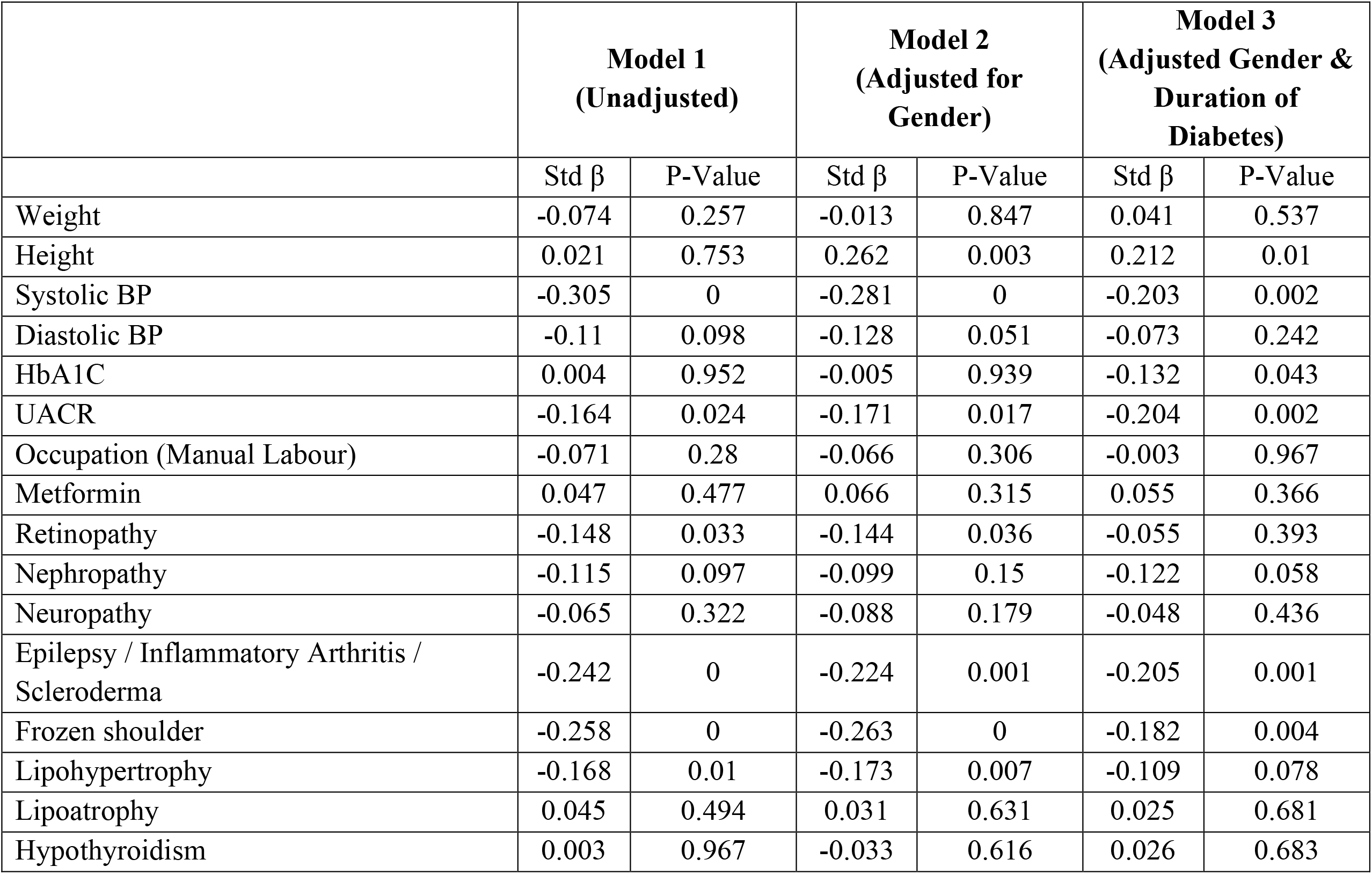
Linear regression for factors contributing to MCP joint limitation

|  | **Model 1**<br>**(Unadjusted)** |  | **Model 2**<br>**(Adjusted for**<br>**Gender)** |  | **Model 3**<br>**(Adjusted Gender &**<br>**Duration of**<br>**Diabetes)** |  |
| --- | --- | --- | --- | --- | --- | --- |
| | Std $\beta$ | P-Value | Std $\beta$ | P-Value | Std $\beta$ | P-Value |
| Weight | -0.074 | 0.257 | -0.013 | 0.847 | 0.041 | 0.537 |
| Height | 0.021 | 0.753 | 0.262 | 0.003 | 0.212 | 0.01 |
| Systolic BP | -0.305 | 0 | -0.281 | 0 | -0.203 | 0.002 |
| Diastolic BP | -0.11 | 0.098 | -0.128 | 0.051 | -0.073 | 0.242 |
| HbA1C | 0.004 | 0.952 | -0.005 | 0.939 | -0.132 | 0.043 |
| UACR | -0.164 | 0.024 | -0.171 | 0.017 | -0.204 | 0.002 |
| Occupation (Manual Labour) | -0.071 | 0.28 | -0.066 | 0.306 | -0.003 | 0.967 |
| Metformin | 0.047 | 0.477 | 0.066 | 0.315 | 0.055 | 0.366 |
| Retinopathy | -0.148 | 0.033 | -0.144 | 0.036 | -0.055 | 0.393 |
| Nephropathy | -0.115 | 0.097 | -0.099 | 0.15 | -0.122 | 0.058 |
| Neuropathy | -0.065 | 0.322 | -0.088 | 0.179 | -0.048 | 0.436 |
| Epilepsy / Inflammatory Arthritis /<br>Scleroderma | -0.242 | 0 | -0.224 | 0.001 | -0.205 | 0.001 |
| Frozen shoulder | -0.258 | 0 | -0.263 | 0 | -0.182 | 0.004 |
| Lipohypertrophy | -0.168 | 0.01 | -0.173 | 0.007 | -0.109 | 0.078 |
| Lipoatrophy | 0.045 | 0.494 | 0.031 | 0.631 | 0.025 | 0.681 |
| Hypothyroidism | 0.003 | 0.967 | -0.033 | 0.616 | 0.026 | 0.683 |

### MRI findings

MRI scans of the hand were performed in 61 patients (12 in <20 degrees mean MCP extension group, 14 in the 20-40 degrees group, 20 in the 40-60 degrees group, and 15 in the >60 degrees group). The radiologist’s findings included flexor tendon thickening or edema in four patients, while one patient has tenosynovial inflammation in the abductor pollicis longus tendon suggestive of De-Quervain tenosynovitis. One patient had an enlarged median nerve suggestive of median nerve neuritis, while one other was found to have a bifid median nerve. Eight patients were found to have ganglion cysts, most commonly in the dorsal scapholunate ligament (n=5), and one had a subcortical cyst. One patient had early degenerative changes in metacarpal joints. All other MRI scans were reported as normal.

When analyzed by group, there was no significant difference within the groups in both mean tendon sheath thickness and maximum tendon sheath thickness.[Table 2] The group with the most stiffness had the highest skin thickness and reduced progressively; but subcutaneous thickness did not differ across groups. Similarly, median nerve area was not different across groups. Both the total hand fibrosis score and the total hand fibrosis Z-score were able to differentiate between the four groups.[Table 2]

Mean palmar skin thickness correlated significantly with palmar fascia thickness (correlation coefficient 0.298, p=0.02) but not with mean tenosynovial thickness (p=0.25). Similarly, mean tenosynovial thickness also correlated with palmar fascia thickness (correlation coefficient 0.28, p=0.03). Univariate linear regression showed that only palmar skin thickness and palmar fascia thickness correlated with MCP angle restriction.[Table 4] Only mean palmar skin thickness remained significantly associated with MCP angle in multiple linear regression.[Table 4]

**Table 4:** Linear regression model for structural contributors to MCP angle

|  | Model 1<br>(Univariate) |  | Model 2 (Multiple<br>univariate) |  |
| --- | --- | --- | --- | --- |
| | Std $\beta$ | P-Value | Std $\beta$ | P-Value |
| Skin Thickness | -0.376 | 0.003 | -0.326 | 0.012 |
| Mean Subcutaneous Thickness | 0.182 | 0.161 | - |  |
| Palmar fascia thickness | -0.262 | 0.041 | -0.165 | 0.193 |
| Median nerve cross sectional area | -0.243 | 0.064 | - |  |
| tendon sheath thickness | -0.032 | 0.808 | - |  |
| R2 | - |  | 13.7% |  |

## Discussion

One third of an Indian cohort of adults with type 1 diabetes had diabetic cheiroarthropathy on clinical evaluation. They were rarely symptomatic or functionally limiting, but did result in limitation of average MCP joint extension. MCP extension limitation had structural correlates on MRI. We found thickened palmar skin in those with MCP extension limitation, while expected features such as inflammatory edema in tendon sheaths or median nerve enlargement were rare. An additive score of tissue thickening was able to differentiate between levels of joint stiffness. These initial data suggest that both the average MCP extension angle in the clinic as well as standardized measurements of palmar structures on hand MRI might help quantify the amount of soft tissue fibrosis in the hand, regardless of the specific diabetes related hand manifestation. Further validation would be important before exploration of associations with other profibrotic manifestations in type 1 diabetes as well as judge response to treatments.

The EDIC/DCCT cohort had nearly double the prevalence of cheiroarthropathy on examination as compared to our cohort; these participants were older (median age 53 years) and consequently had had a much longer diabetes duration (32 years).[2] A questionnaire-based evaluation in a Danish patient registry, (n=2200) also reported a 66% prevalence of upper extremity impairments, although these also included adhesive capsulitis. Studies show a decline in these manifestations over time, and LJM reduced from 43 to 23% in adult patients with type 1 diabetes over two decades.[7] Reduced prevalence has been attributed to better glycemic care, however the association of hand manifestations with HbA1C are inconsistent.[20] Associations with microvascular disease including retinopathy and nephropathy have been reported [2, 21] while others have failed to replicate these.[13] We found an association of the MCP angle extension with renal involvement alone, and similar associations were found with MCP and wrist flexion angles.[22] Across studies and methods of assessment, age and duration of diabetes are pervasively associated with developing cheiroarthropathy.[2, 7, 13]

Despite the joint stiffness seen on clinical examination, these conditions were neither commonly symptomatic, nor functionally limiting. While the DHI was higher in those with more severe joint restriction, differences would not be considered clinically significant.[19] Other functional scores used in other studies echo the insubstantial functional impact of these conditions: Health assessment questionnaire (HAQ) scores in patients with DD were increased by a 0.15 as compared to controls.[3] The DASH disability score of nine seen in the EDIC cohort was also less than the minimally clinically important score of 10.8.[2, 23] In a large Taiwanese study, only about 10% of patients with diabetes volunteered symptoms in the hand.[8] This wide discrepancy between the prevalence of patient-volunteered symptoms and physician-examined manifestations suggest an insidiously developing, non-inflammatory trajectory of soft tissue fibrosis.

Patients who are symptomatic for diabetic cheiroarthropathy are likely to represent a subset with advanced stiffness; a consistent, reproducible, and quantitative measure of hand stiffness would be useful in detecting early manifestations. The prayer sign and the tabletop test, both traditionally used for the diagnosis, are non-quantitative. Goniometry has been used to show that patients with diabetes have stiffer joints.[24] This analysis showed that bar a few small joints, all joints assessed were less flexible in the diabetic population, although a stepwise regression failed to identify a single ‘sentinel’ joint. However, the MCP and distal interphalangeal (DIP) joints were most severely affected. Since all described manifestations within the cheiroarthropathy umbrella result in preferential fibrotic thickening of one or more soft tissue structures on the flexor aspect of the hand, we selected resistance to MCP stretch as measured by maximum passive extension as a metric that would intuitively encapsulate stiffness regardless of the cause. While LJM commonly affects all fingers, DD commonly affects the fourth and fifth digits, and flexor tenosynovitis may affect one or multiple digits. Unlike the table top test, an average of individual MCP extensions would pick up all of these. Indeed, in our study, those with a physician-diagnosed cheiroarthropathy had a much lower possible MCP extension on average than those who did not, with a clinically significant difference of nearly thirty degrees. On the contrary, flexion is relatively preserved; reflected in the grip strength not being significantly affected. While we did not check for MCP flexion, it is likely to be far less affected; average MCP flexion was more than 80 degrees in a Type 1 longitudinal cohort, and patients had an approximate loss of 10 degrees over 15 years.[22]

We found that hand stiffness, as evidenced by restriction of MCP extension, had imaging correlates on MRI. Previous MRI descriptions are few and are limited to a single case report. Khanna and Ferguson reported synovial sheath thickening and edema along all flexor sheaths on Coronal T2-weighted images, and axial T1 fat-saturation gadolinium enhanced images showed tenosynovial proliferation.[17] This patient likely had flexor tenosynovitis; however, other conditions under the diabetic cheiroarthropathy umbrella such as LJM and DD were not represented. Flexor tendon sheath and subcutaneous tissue thickening were found on ultrasound in diabetic cheiroarthropathy.[25] All these published descriptions have thus far been subjective, and in patients who had severe, diagnosed manifestations. With only qualitative descriptions available, it is difficult to quantitate mobility limitation as well as judge disease progression. We found that an intuitive MRI score that includes skin, tendon and palmar fascia thickening could differentiate between patients with and without hand stiffness. Further work would be needed to validate its utility in larger datasets, judge reproducibility and sensitivity to change.

We found that skin thickening contributed most to limited joint mobility in this group, while subcutaneous tissue thickening did not play as much of a role. This finding contrasts to both the US findings, and may arise not only from differences in diagnostic modality, the heterogeneous distribution of subcutaneous fat in the hand, and the inclusion of patients with joint mobility limitation regardless of cause.[25] Calculating subcutaneous fat volumes might help understand this discrepancy. Another striking feature in this cohort was the relatively normal size of the median nerve, even in those patients with symptoms suggestive of neuropathy, albeit, these symptoms were not confirmed on nerve conduction. This observation may suggest that chronic, compressive neuropathy in type 1 diabetes may not have the expected increase in median nerve cross sectional area routinely used to diagnose CTS.[26] This finding supports other studies in which the median nerve cross sectional area was smaller in diabetics with CTS (mean 8.8 mm^2^) than with patients with CTS without diabetes (mean 10.4 mm^2^).[27] More work in this area would be useful in determining if patients with diabetes need separate cut-off values in diagnosing CTS.

Regardless of the degree of hand stiffness, inflammation on MRI as evidenced by tissue edema was conspicuous by its absence. However, gadolinium contrast was not used, which might have potentially highlighted inflammation. This finding contrasts with the MRI findings in autoimmune conditions associated with flexor tenosynovitis such as rheumatoid arthritis [28] and systemic sclerosis.[29] Tissue edema was seen in tenosynovial sheaths in the earlier MRI description of diabetic cheiroarthropathy [17] and in a small number of our patients as well. It is possible that the time of assessment along the temporal course of the manifestation makes a difference, and that these conditions are inflammatory to start with, with fibrosis coming later. However, most inflammatory tenosynovitis cases are painful, while pain was rare in our group.[28] It could be postulated that pathways to fibrosis are therefore distinct and AGE deposition, hypoxia etc. might be more important disease mechanisms than inflammation.

This study has many strengths: to our knowledge, it is the first systematic evaluation of joint stiffness in type 1 diabetes using MRI imaging, and the structural differences in skin thickness provide credence to the utility of the MCP extension angle as a quantifiable clinical measure of hand fibrosis. We did not restrict to a single hand manifestation but combined them under a common, pathogenesis-backed umbrella. Both clinical measurements and MRI measurements were performed by more than one assessor, suggesting a reproducibility of methods. With these initial descriptions of methods of quantification of fibrosis load, we lay grounds to further exploration of granular associations with internal organ fibrosis signals with hand fibrosis, as well as possible clinical outcome measures for treatment modalities in diabetic cheiroarthropathy. This study has limitations: even the ‘controls’ with no hand stiffness were diabetic, and future work would include age-matched non-diabetic controls. We did not compare the performance of MCP extension to other joints. The groups of 20 degrees were arbitrarily decided based on perceived convenience for a clinician. A goniometer was not used to measure MCP extension angles; however, the lack of specialized equipment would, in our opinion, make the screening measurement easier to perform in the community.

In conclusion, we found a substantial subset of patients with adult type 1 diabetes had diabetic cheiroarthropathy and resulting limitations in MCP extension. Joint stiffness was driven by skin thickening, and inflammatory tenosynovial involvement was rare on MRI. Joint mobility limitation was rarely symptomatic or functionally limiting, suggesting that an active search in the clinic would be warranted. Our data suggest that average MCP joint extension in the clinic, and measuring thicknesses of the skin, palmar fascia, and tendon sheaths on MRI, could serve as measures of cumulative fibrosis in the hand. Pending more extensive validation and sensitivity to change, these measurements could potentially be used to elucidate associations with internal organ fibrosis as well as outcome measures for trials for musculoskeletal fibrosis.

## Data Availability

All clinical and imaging data are stored at the Diabetes Unit, KEM Hospital Research Centre.

## Data sharing

All data including MRI scans is stored at the Diabetes Unit, KEM Hospital Research Centre. Data sharing requests can be made to Dr Sanat Phatak with an analysis plan.

## Acknowledgements and affiliations

Authors thank Prof Satyajit Rath for methodological guidance; Dr Kalpana Jog, Vidya Gokhale, Swati Alekar, Dr Neelima Nagarkar from the type 1 diabetes clinic; Rasika Ladkat for study related administration. This study was funded via a DBT/Wellcome India Alliance Fellowship to Dr Sanat Phatak.

## References

1. Rosenbloom AL, Frias JL. Diabetes mellitus, short stature and joint stiffness - a new syndrome. Clin Res. 1974;22:92A.

2. Larkin ME, Barnie A, Braffett BH, Cleary PA, Diminick L, Harth J, et al. Musculoskeletal complications in type 1 diabetes. Diabetes Care. 2014 Jul;37(7):1863–9.

3. Gutefeldt K, Lundstedt S, Thyberg ISM, Bachrach-Lindström M, Arnqvist HJ, Spångeus A. Clinical Examination and Self-Reported Upper Extremity Impairments in Patients with Long-Standing Type 1 Diabetes Mellitus. J Diabetes Res. 2020;2020:4172635.

4. Rydberg M, Zimmerman M, Gottsäter A, Svensson AM, Eeg-Olofsson K, Dahlin LB. Diabetic hand: prevalence and incidence of diabetic hand problems using data from 1.1 million inhabitants in southern Sweden. BMJ Open Diabetes Res Care. 2022 Jan;10(1):e002614.

5. Cagliero E. Rheumatic manifestations of diabetes mellitus. Curr Rheumatol Rep. 2003 Jun;5(3):189–94.

6. Pandey A, Usman K, Reddy H, Gutch M, Jain N, Qidwai S. Prevalence of hand disorders in type 2 diabetes mellitus and its correlation with microvascular complications. Ann Med Health Sci Res. 2013 Jul;3(3):349–54.

7. Lindsay JR, Kennedy L, Atkinson AB, Bell PM, Carson DJ, McCance DR, et al. Reduced prevalence of limited joint mobility in type 1 diabetes in a U.K. clinic population over a 20-year period. Diabetes Care. 2005 Mar;28(3):658–61.

8. Chen LH, Li CY, Kuo LC, Wang LY, Kuo KN, Jou IM, et al. Risk of Hand Syndromes in Patients With Diabetes Mellitus: A Population-Based Cohort Study in Taiwan. Medicine (Baltimore). 2015 Oct;94(41):e1575.

9. Kameyama M, Chen KR, Mukai K, Shimada A, Atsumi Y, Yanagimoto S. Histopathological characteristics of stenosing flexor tenosynovitis in diabetic patients and possible associations with diabetes-related variables. J Hand Surg Am. 2013 Jul;38(7):1331–9.

10. Gokcen N, Cetinkaya Altuntas S, Coskun Benlidayi I, Sert M, Nazlican E, Sarpel T. An overlooked rheumatologic manifestation of diabetes: diabetic cheiroarthropathy. Clin Rheumatol. 2019 Mar;38(3):927–32.

11. Merashli M, Chowdhury TA, Jawad AS. Musculoskeletal manifestations of diabetes mellitus. QJM. 2015 Nov;108(11):853–7.

12. Casanova JE, Casanova JS, Young MJ. Hand function in patients with diabetes mellitus. South Med J. 1991 Sep;84(9):1111–3.

13. Frost D, Beischer W. Limited joint mobility in type 1 diabetic patients: associations with microangiopathy and subclinical macroangiopathy are different in men and women. Diabetes Care. 2001 Jan;24(1):95–9.

14. Broekstra DC, Groen H, Molenkamp S, Werker PMN, van den Heuvel ER. A Systematic Review and Meta-Analysis on the Strength and Consistency of the Associations between Dupuytren Disease and Diabetes Mellitus, Liver Disease, and Epilepsy. Plast Reconstr Surg. 2018 Mar;141(3):367e–379e.

15. Uiterwaal CS, Grobbee DE, Sakkers RJ, Helders PJ, Bank RA, Engelbert RH. A relation between blood pressure and stiffness of joints and skin. Epidemiology. 2003 Mar;14(2):223–7.

16. Ban CR, Twigg SM. Fibrosis in diabetes complications: pathogenic mechanisms and circulating and urinary markers. Vasc Health Risk Manag. 2008;4(3):575–96.

17. Khanna G, Ferguson P. MRI of diabetic cheiroarthropathy. AJR Am J Roentgenol. 2007 Jan;188(1):W94–5.

18. Maran A, Morieri ML, Falaguasta D, Avogaro A, Fadini GP. The Fast-Glycator Phenotype, Skin Advanced Glycation End Products, and Complication Burden Among People With Type 1 Diabetes. Diabetes Care. 2022 Oct 1;45(10):2439–44.

19. Turan Y, Duruöz MT, Aksakalli E, Gürgan A. Validation of Duruöz Hand Index for diabetic hand dysfunction. J Investig Med. 2009 Dec;57(8):887–91.

20. McCance DR, Crowe G, Quinn MJ, Smye M, Kennedy L. Incidence of microvascular complications in type 1 diabetic subjects with limited joint mobility: a 10-year prospective study. Diabet Med. 1993 Nov;10(9):807–10.

21. Garg SK, Chase HP, Marshall G, Jackson WE, Holmes D, Hoops S, et al. Limited joint mobility in subjects with insulin dependent diabetes mellitus: relationship with eye and kidney complications. Arch Dis Child. 1992 Jan;67(1):96–9.

22. Labad J, Rozadilla A, Garcia-Sancho P, Nolla JM, Montanya E. Limited Joint Mobility Progression in Type 1 Diabetes: A 15-Year Follow-Up Study. Int J Endocrinol. 2018;2018:1897058.

23. Franchignoni F, Vercelli S, Giordano A, Sartorio F, Bravini E, Ferriero G. Minimal clinically important difference of the disabilities of the arm, shoulder and hand outcome measure (DASH) and its shortened version (QuickDASH). J Orthop Sports Phys Ther. 2014 Jan;44(1):30–9.

24. Schulte L, Roberts MS, Zimmerman C, Ketler J, Simon LS. A quantitative assessment of limited joint mobility in patients with diabetes. Goniometric analysis of upper extremity passive range of motion. Arthritis Rheum. 1993 Oct;36(10):1429–43.

25. Ismail AA, Dasgupta B, Tanqueray AB, Hamblin JJ. Ultrasonographic features of diabetic cheiroarthropathy. Br J Rheumatol. 1996 Jul;35(7):676–9.

26. Moran L, Perez M, Esteban A, Bellon J, Arranz B, del Cerro M. Sonographic measurement of cross-sectional area of the median nerve in the diagnosis of carpal tunnel syndrome: correlation with nerve conduction studies. J Clin Ultrasound. 2009;37(3):125–31.

27. Guillen-Astete CA, Luque-Alarcon M, Garcia-Montes N. Ultrasound Assessment of the Median Nerve Does Not Adequately Discriminate the Carpal Tunnel Syndrome among Patients Diagnosed with Diabetes. Diabetology. 2021 Nov 1;2(4):226–31.

28. Eshed I, Feist E, Althoff CE, Hamm B, Konen E, Burmester GR, et al. Tenosynovitis of the flexor tendons of the hand detected by MRI: an early indicator of rheumatoid arthritis. Rheumatology (Oxford). 2009 Aug;48(8):887–91.

29. Low AH, Lax M, Johnson SR, Lee P. Magnetic resonance imaging of the hand in systemic sclerosis. J Rheumatol. 2009 May;36(5):961–4.

